## Supplementary material for "The impact of antenatal balanced plate nutrition education for pregnant women on birth weight: a cluster randomised controlled trial in rural Bangladesh": S1 Table

**S1 Table: The mean birth weight and the adjusted mean difference in birth weight of balanced -plate intervention compared to usual programs.**

|  | **Number of participants** | | **Birth weight (g)** | | | | **Adjusted mean difference^a^** | | | |
| --- | --- | --- | --- | --- | --- | --- | --- | --- | --- | --- |
|  | **Intervention** | **Control** | **Intervention**  Mean (SD) | | **Control**  Mean (SD) | | **Estimate** | **(95%CI)** | **P-value** | **P-value for interaction** |
| **Overall** | 382 | 397 | 2861.0 | (444.2) | 2736.8 | (423.2) | 121.8 | (11.7, 231.9) | 0.030 |  |
| **Mother’s age^1^ (years)** |  |  |  |  |  |  |  |  |  | 0.005 |
| <20 | 80 | 64 | 2916.3 | (471.1) | 2637.5 | (481.0) | 290.6 | (111.9, 469.3) | 0.001 |  |
| >=20 | 288 | 313 | 2847.6 | (443.4) | 2762.6 | (401.1) | 78.8 | (-26.7, 184.3) | 0.143 |  |
| **Mother’s education^2^** |  |  |  |  |  |  |  |  |  | 0.237 |
| None | 82 | 89 | 2869.5 | (460.6) | 2693.3 | (339.7) | 129 | (6.1, 251.9) | 0.021 |  |
| Primary | 250 | 242 | 2839.2 | (443.9) | 2761.6 | (394.4) | 75.7 | (-37.4, 188.9) | 0.190 |  |
| Secondary or higher | 40 | 55 | 2982.5 | (435.5) | 2741.8 | (588.0) | 204.3 | (-34.8, 443.5) | 0.094 |  |
| **Family income^3^** |  |  |  |  |  |  |  |  |  | 0.1663 |
| Low | 87 | 79 | 2819.5 | (397.9) | 2663.3 | (420.4) | 217.8 | (60.9, 374.6) | 0.007 |  |
| Medium | 190 | 209 | 2806.8 | (421.7) | 2750.7 | (390.8) | 37.3 | (-71.7, 146.3) | 0.503 |  |
| High | 87 | 84 | 2995.4 | (514.5) | 2791.7 | (433.2) | 177 | (-40.7, 394.7) | 0.111 |  |

Birth weight was analyzed using linear regression. ^a^ We adjusted all models for antenatal care from SK, antenatal care from health centers, use of iron and folic acid and clusters using generalized estimating equation (GEE) models assuming exchangeable correlation structure and applied sandwich estimator to standard errors.. ^1^ n=745 due to 34 missing maternal age values. ^2^ n=758 due to 21 missing maternal education values. ^3^ n=736 due to 43 missing family income values.
