## Supplementary material for "The impact of antenatal balanced plate nutrition education for pregnant women on birth weight: a cluster randomised controlled trial in rural Bangladesh": S2 Table

**S2 Table: Percentage of low birth weight and adjusted relative risk of balanced-plate intervention compared to usual programs.**

|  | **Number of participants** | | **Low birth weight (<2,500g)** | | | | **Adjusted relative risk^a^** | | | |
| --- | --- | --- | --- | --- | --- | --- | --- | --- | --- | --- |
|  | **Intervention** | **Control** | **Intervention**  **n (%)** | | **Control**  **n (%)** | | **Estimate** | **(95%CI)** | **P-value** | **P-value for interaction** |
| **Overall** | 382 | 397 | 37 | (10) | 87 | (22) | 0.53 | (0.29, 0.98) | 0.042 |  |
| **Mother’s age^1^ (years)** |  |  |  |  |  |  |  |  | 0.183 | 0.183 |
| <20 | 80 | 64 | 6 | (8) | 17 | (27) | 0.29 | (0.13, 0.68) | 0.004 |  |
| >=20 | 288 | 313 | 31 | (11) | 62 | (20) | 0.58 | (0.32, 1.04) | 0.068 |  |
| **Mother’s education^2^** |  |  |  |  |  |  |  |  | 0.262 | 0.262 |
| None | 82 | 89 | 11 | (13) | 22 | (25) | 0.55 | (0.29, 1.05) | 0.071 |  |
| Primary | 250 | 242 | 25 | (10) | 47 | (19) | 0.55 | (0.3, 1) | 0.049 |  |
| Secondary or higher | 40 | 55 | 1 | (3) | 12 | (22) | 0.10 | (0.02, 0.7) | 0.020 |  |
| **Family income^3^** |  |  |  |  |  |  |  |  | 0.362 | 0.362 |
| Low | 87 | 79 | 9 | (10) | 25 | (32) | 0.31 | (0.13, 0.71) | 0.006 |  |
| Medium | 190 | 209 | 23 | (12) | 41 | (20) | 0.66 | (0.36, 1.2) | 0.170 |  |
| High | 87 | 84 | 5 | (6) | 12 | (14) | 0.43 | (0.14, 1.35) | 0.149 |  |

Low birth weight (LBW) was analyzed using binomial regression with a log link. ^a^ We adjusted all models for antenatal care from SK, antenatal care from health centers, use of iron and folic acid and clusters using generalized estimating equation (GEE) models assuming exchangeable correlation structure and applied sandwich estimator to standard errors. ^1^ n=745 due to 34 missing maternal age values. ^2^ n=758 due to 21 missing maternal education values. ^3^ n=736 due to 43 missing family income values.
